# The benefits of palliative care follow-up combining day hospital and telemedicine

**DOI:** 10.1101/2025.01.12.25320435

**Authors:** Valerie Mauries Saffon, Alfonsina Faya-Robles, Marie Bourgouin, Sebastien Lamy, Nathalie Caunes-Hilary, Bettina Couderc

**Affiliations:** Department of palliative care, Claudius Regaud-Oncopole, 1 avenue Irène Joliot Curie 31059 Toulouse cedex 9, France; Regard Social (Association loi 1901 - Sciences Humaines et Sociales, Santé, sociétés), 5 rue de Kiev, Maison Ouverte de Santé, 31100 Toulouse, France; Department Equity, CERPOP, UMR 1295, Inserm - Université Paul Sabatier -Toulouse III : “Trajectoires d’innovations en santé : enjeux bioéthiques et sociétaux” https://cerpop.inserm.fr 37 allées Jules Guesde, 31073 Toulouse cedex, France; Department Bioethics, CERPOP, UMR 1295, Inserm - Université Paul Sabatier -Toulouse III : “Trajectoires d’innovations en santé : enjeux bioéthiques et sociétaux” https://cerpop.inserm.fr, 37 allées Jules Guesde, 31073 Toulouse cedex, France

## Abstract

The objective of the present study is to evaluate the impact of remote consultations (telemedicine) conducted on a monthly basis, in conjunction with quarterly inpatient follow-ups conducted by a team specialising in palliative care for cancer patients, in terms of anticipating potential deterioration in patients’ general condition and enhancing their quality of life. The study also investigates whether there is a specific profile of patients who could benefit from teleconsultation and identifies patients’ expectations and concerns regarding teleconsultation.

The study enrolled 36 patients on their initial admission to a palliative care day hospital (HDJ), and they completed a questionnaire regarding their demographics, expectations, and reservations concerning teleconsultation. Patients were then scheduled for two monthly teleconsultations and were advised to return to the HDJ at D = 3 months. However, only 13 out of the 36 patients enrolled were able to benefit from the two teleconsultations and return to the HDJ after 3 months. This finding underscores the critical issue of patients being referred to the mobile palliative care team at a late stage. Our study revealed that factors such as age, gender, socio-economic status, and pathology did not influence patient compliance or the efficacy of the teleconsultations. The study emphasised the benefits of monthly teleconsultations for the clinical and psychological management of patients, highlighting that patients had minimal negative preconceptions upon enrolment and expressed a strong desire to continue these monthly consultations after the initial 3-month period. The findings underscore the significance of leveraging teleconsultations for patients receiving palliative care in oncology, particularly in scenarios where face-to-face consultations are not feasible. Telemedicine holds immense potential to enhance the monitoring of a larger patient population, thereby promoting greater equity in healthcare delivery.

## INTRODUCTION

Recent years have seen a steady increase in life expectancy for cancer patients, attributable to advancements in treatment and care, including therapeutic innovations, targeted therapies and immunotherapies, as well as early diagnosis [1][2]. While the probability of a cancer patient being cured varies according to factors such as geographical location, histological type, stage at diagnosis, country of residence and age, it is estimated that between 21% and 47% of male patients, and between 38% and 59% of female patients, have the potential to be cured of their cancer [3]. While these figures are encouraging, they underscore the pivotal role of the palliative care physician in oncology care, as a substantial proportion of patients will experience disease stabilisation or progression and necessitate specialised monitoring [4,5,6]. In the absence of a curative plan, it is imperative to provide appropriate care and treatment that is not only aimed at controlling symptoms (e.g. pain and anxiety) but also at ensuring a good quality of life for patients with a chronic disease. In recent years, there has been an increasing scarcity of palliative care beds [9–11].

The paucity of palliative care day hospital and long-term hospital places has prompted government authorities to consider how to improve care in this area, whether by creating additional beds, new palliative care teams or new types of care. Teleconsultation, i.e. the use of telecommunication technologies to provide remote medical consultations, has emerged as a potential method for improving care in general and can be proposed for the monitoring of palliative care patients. This method involves a secure remote consultation between a patient and their doctor, facilitated by a computer, tablet or smartphone. The concept of teleconsultation was introduced in France in 2009, with subsequent legislative codification in 2018. [12]

While telemedicine cannot substitute for face-to-face consultations, [13] it can enhance care by enabling timely consultations for patients while they await physical attendance at a care facility. The efficacy of teleconsultation in improving the management of symptoms and the quality of life for palliative care patients has been demonstrated by continuous monitoring and timely interventions. Furthermore, it has the potential to reduce the number of patients requiring hospitalisation in emergency situations between consultations.

The existing body of literature on the establishment of teleconsultation in palliative care indicates that the implementation of such consultations tends to yield high levels of satisfaction among patients, carers and home care providers [14]. This positive evaluation of teleconsultation is predicated on several observations. Firstly, a randomised clinical trial demonstrated that telemedicine facilitates direct, patient-centred communication between palliative care clinicians, patients, surrogates and carers in the home, thereby enhancing collaboration and integration of care [15]. The involvement of a home care worker (nurse) in the consultation with the patient has also been identified as a key benefit, facilitating swift referrals to general practitioners [16–18]. The enhanced communication between hospital and home care providers has been demonstrated to alleviate the burden on caregivers. This is achieved by eliminating the need for frequent patient transport to medical appointments, thereby reducing the emotional strain associated with informing family members of treatment modifications [19].Moreover, the incorporation of monthly teleconsultations has been shown to diminish the burden on families by providing prompt and accessible support, consequently reducing the reliance on emergency services [20]. Teleconsultation facilitates better integration of primary and specialist palliative care, leading to improved continuity of care and collaborative treatment planning [16,17]. In terms of quality of care, several studies show that telemedicine promotes coordination between home care and hospital-based specialists, enabling patients to receive comprehensive care tailored to their needs [16,17][21]. Whilst some studies suggest that frequent teleconsultations may lead to an increase in symptom burden due to increased attention to symptoms and potential distress [22], the majority of studies highlight that continuous monitoring leading to timely interventions through telemedicine improves symptom management and quality of life for palliative care patients [20,23][19].For carers, teleconsultation saves clinicians time, reduces waiting times for appointments and reduces absenteeism, making it an effective method of delivering palliative care. It also helps to reduce unnecessary admissions and transfers to hospital, thereby reducing the overall burden on healthcare systems [23][20].

It has been hypothesised that the implementation of teleconsultation could serve to reduce health inequalities by extending healthcare access to a greater number of patients. However, it is imperative to acknowledge the challenges associated with the cultural and social context of healthcare, which necessitates a thorough examination of the roles and responsibilities of legislators, government representatives, and health care providers [21] with regard to telemedicine. In addition to the geographical parameters associated with the physical environment of patients, other dimensions related to socio-economic categories may constitute a difficulty of access for patients (lack of material and digital resources and of suitable accommodation for carrying out teleconsultations).As discussed in the literature on access to health care [24,25], it is no longer sufficient to reduce health inequalities to a question of spatial or geographical accessibility. Other equally important dimensions must be considered, including the availability of health services and professionals (e.g. waiting times, consultation duration, specialties practised), the convenience and adaptation to people’s needs and constraints (e.g. hours), the financial and material capacity of patients (e.g. transportation for face-to-face consultations or technical equipment for teleconsultations), and the quality of the information and procedures co-produced during the consultation. In this context, the research questions focused on the potential benefits of teleconsultation for specific age, gender or socio-professional categories, particularly in the context of supportive care in oncology. The objective was to transition from a binary (yes/no) response framework and to explore the adaptability of this consultation modality to the unique characteristics of the service [26]. In addition, the focus was on access to care, defined not in geographical but in social terms, and in particular the risk of reproducing social inequalities in health. In other words, the objective was to determine whether there is a typical patient profile for whom teleconsultation is beneficial. In addressing this question, we sought to explore a patient profile for whom telemedicine is advantageous, adopting a perspective that diverges from the concept of ’acceptability’. Instead, we emphasised the conception of telemedicine as a malleable instrument, adaptable to various factors, including the characteristics of the patient, the expertise of the professional, and the dynamics of the therapeutic relationship, among other considerations.

## MATERIALS AND METHODS

Patient recruitment: Patients attending a palliative day hospital for the first time were offered inclusion in the research protocol. This day hospital (HDJ) generally allows for a global assessment of the patient (physical, psychosocial), the setting up of home help, regular follow-up, reflection on future care (oncological and symptomatic, thanks to interdisciplinary reflection), ethical questions; sometimes it intervenes at the time of a refusal of care on behalf of the patient or a request for euthanasia.

The inclusion criteria encompassed cancer patients who were referred to the mobile palliative care team (EMSP) for the first time and who possessed a smartphone or a connected computer/tablet, with a life expectancy of 3 months or more. At the commencement of the day, the patients were provided with an information leaflet, and at the conclusion of the day, the investigator addressed their queries regarding the research and obtained their written consent.

The patients then proceeded to complete a demographic questionnaire and responded to questions concerning their expectations, hopes and fears regarding teleconsultation.

The present questionnaire was utilised for the purpose of conducting a descriptive quantitative analysis of the characteristics of the respondents.

The schedule of teleconsultations and day hospital appointments was established.

<u>Teleconsultations:</u> Patients received an e-mail three days prior to their scheduled teleconsultation appointment, containing a link to access the consultation. On the day of the consultation, the EMSP doctor who had previously met the patient logged on to conduct an overall assessment of the patient, in accordance with the HDJ protocol. This teleconsultation enabled the exploration of any discomfort or symptoms, and the assessment of the patient’s autonomy, nutritional status, and psychological well-being. During each session, patients were prompted to disclose the presence or absence of domestic assistance, particularly the presence of a carer, and any social challenges they might be facing. Often, the patient would join the session with the primary carer, providing an opportunity for the medical team to establish a rapport with them. The camera was utilized to assess the general condition of both the patient and the carer, encompassing aspects such as pain, weight loss, asthenia, and independence. The camera was also utilized to visualize the location of specific symptoms (pain) by the patient in front of the camera. Finally, the patient was asked about their current and future life plans. At the conclusion of the teleconsultation, a summary was prepared, including, if necessary, a prescription for treatment (prescription sent by e-mail to the patient or to the pharmacy).The patient was informed of the next teleconsultation or day clinic follow-up appointment.

The link with the patient’s general practitioner or care structure has been established.

HDJ visit at month 3: A face-to-face reassessment was carried out in the palliative care HDJ, utilizing the same procedures as the initial assessment; this evaluation enabled a comprehensive clinical examination to be conducted. During this HDJ, a semi-structured interview was conducted with the patient to assess their sentiments regarding the teleconsultations they had received. This interview was recorded and subsequently transcribed for a qualitative study (sociological thematic analysis) of thirteen interviews with patients included in the study. The method of analysis used was manual and thematic, “i.e. the transformation of a given corpus into a certain number of themes representative of the content analyzed, in relation to the research orientation (the problem)” [27].

The study received approval from the Research Ethics Committee of the Université Fédérale de Toulouse on 17 February 2022 (approval number 2022-473).

The study began (inclusion of the first patient) on 10 March 2022 and ended on 1 July 2024.

## RESULTS

### Demographics

The participants of the study are displayed in Table 1. The study was offered to 38 patients, of whom 2 were excluded due to their inability to communicate. The final study population comprised 29 female and 7 male patients. The median age of the patients was 63.5 years, with 50% of patients falling between the ages of 52 and 72 years. The majority of participants (86%) did not live alone, either as a couple or with a close relative. The geographical distribution of patients revealed that 53% resided more than 50 km from the care center. A significant proportion, 80%, possessed an educational attainment that was equal to or higher than the French baccalauréat. Furthermore, 72% of patients reported having a comfortable standard of living, with no patients included in the study who exhibited a precarious standard of living. It is noteworthy that no exclusion criteria related to social status were identified. The study also examined the patients’ smartphone (or tablet/computer) usage patterns, with all participants reporting that they consulted their device at least once a day. Furthermore, 20% of participants had previously engaged in teleconsultation, while 100% of patients who were offered participation and had access to a connection agreed to be included in the study. Patients’ feelings about telemedicine before their first consultation.

Following enrolment in the study, patients completed a questionnaire on their perceptions, expectations and/or fears regarding telemedicine in general and their own follow-up on the day of their first consultation with the EMSP. The aim of this questionnaire was to assess patients’ general and personal understanding of telemedicine. The first observation to be made is that all patients who were offered monthly telemedicine follow-up as an alternative to exclusive inpatient follow-up every three months accepted. The rationale behind patient acceptance of the proposed changes was investigated, and the results indicated that for 100% of patients, the primary motivators were the aspiration for enhanced follow-up (monthly in lieu of quarterly) and the assurance of increased accessibility to medical consultations (quality of follow-up). The feasibility of providing monthly face-to-face consultations was not a subject of inquiry, as such an option was not available. However, 94% of patients responded in the affirmative to the question, “I think telemedicine saves me having to come to the clinic to solve my medical problems” (Fig 1A). This finding indicates a high level of confidence in the benefits of teleconsultation.

**Figure 1:**
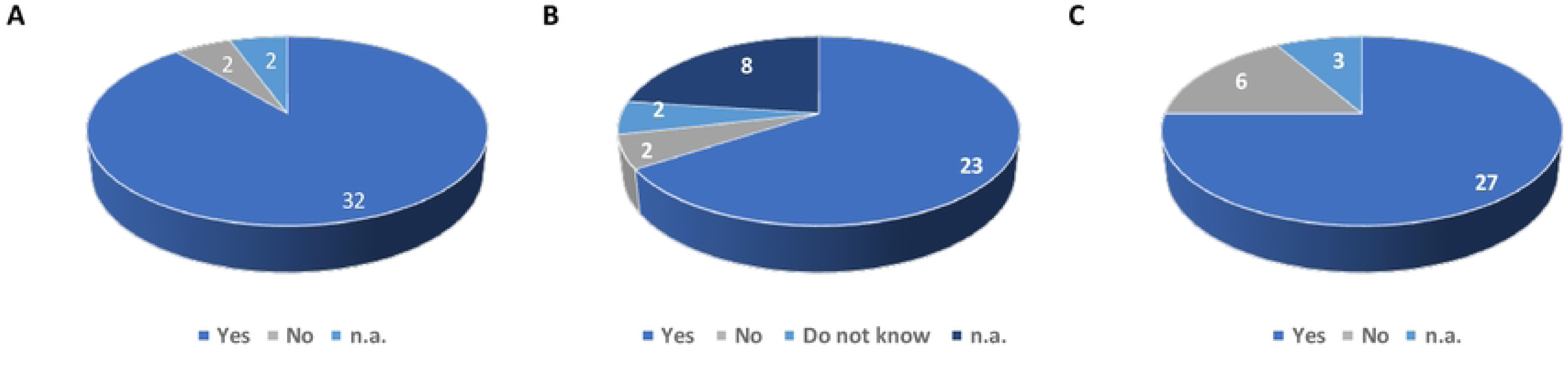
Theoretical advantages of teleconsultation for patients newly admitted to palliative care A: Patient responses to the item: “I believe that telemedicine eliminates my need to visit the medical facility for my health-related inquiries.” B: Patient responses to the item: “Telemedicine reduces my time expenditure.” C: Patient responses to the item: Telemedicine will make it easier for my family and friends to accompany me to the consultation compared to day hospitalisation. n.a. not applicable (patients did not answer the question)

A survey of patients revealed that 85% of respondents expressed the belief that teleconsultation would result in a reduction in their fatigue levels (Fig. 1B). Additionally, 82% of respondents indicated that such consultations would be less disruptive to those around them (Fig. 1C), emphasizing the convenience aspect. With respect to the perceived limitations of telemedicine, patient opinions were more divided on the relevance of teleconsultations in addressing all their queries and ensuring optimal medical follow-up (Fig. 2). In response to the question, “I think telemedicine could be useful for me, but only for certain medical procedures,” 94% of respondents answered in the affirmative (Fig. 2A). It is noteworthy that 57% of respondents did not consider telemedicine to be suitable for their medical follow-up (Fig. 2B), whereas 50% of respondents believed that teleconsultation would be as beneficial as a face-to-face consultation. This finding suggests a degree of mistrust regarding the effectiveness of teleconsultation in terms of diagnosis and treatment, despite the absence of first-hand experience.

**Figure 2:**
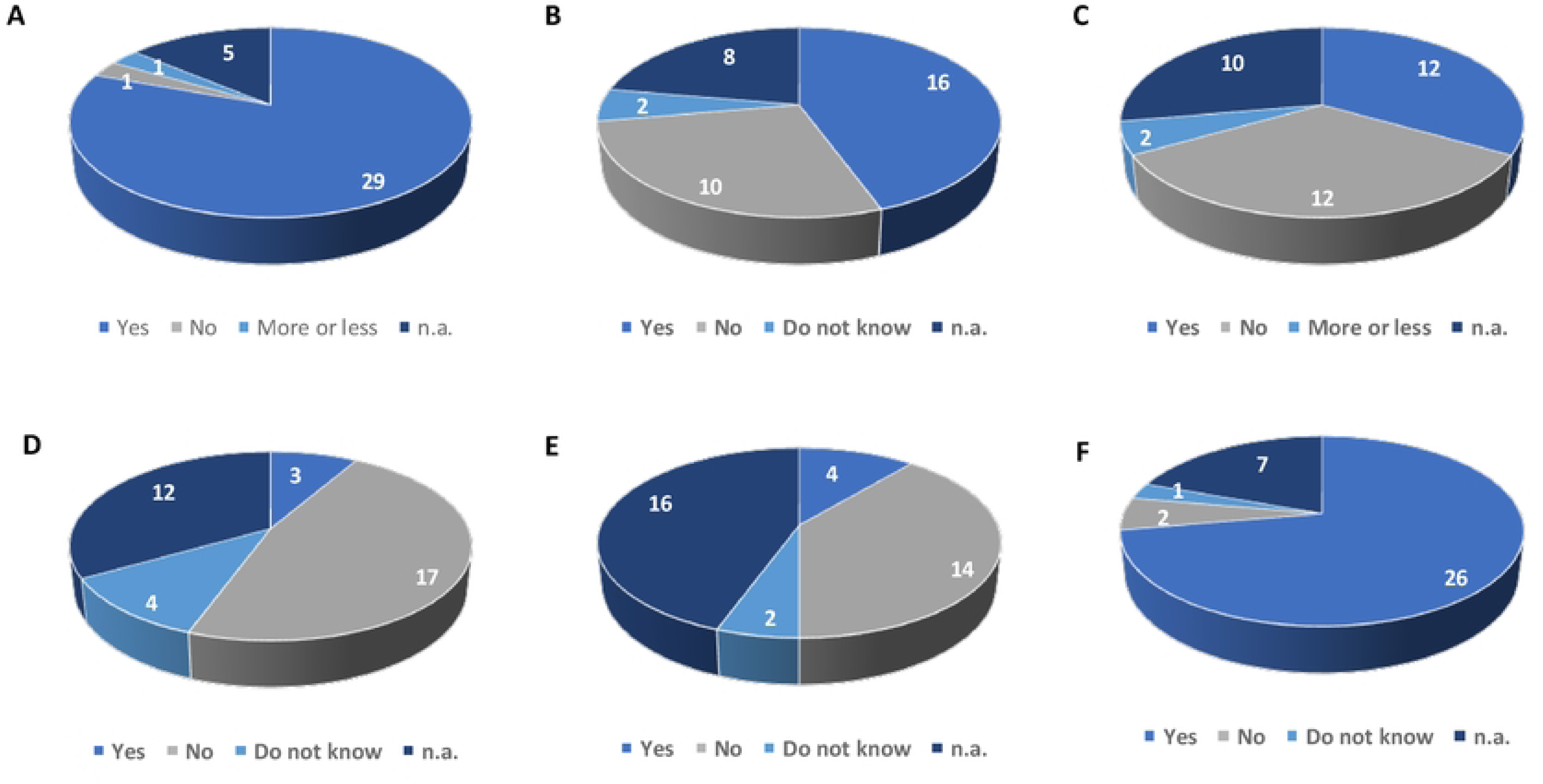
Believed limitations of tele-consultation for patients newly involved in palliative care A: Responses of the patients to the item ’I think telemedicine could be useful for me, but only for certain medical procedures’. B: Responses of the patients to the item: ’I don’t think telemedicine is suitable for my medical follow-up’. C: Responses of the patients to the item: ’A telemedicine consultation is just as useful as a face-to-face consultation’. D: Responses of the patients to the item: ’Telemedicine dehumanises medicine’. E: Responses of the patients to the item: ’Telemedicine weakens relationships with carers’. F: Responses of the patients to the item: ’Monthly telemedicine helps to maintain a strong link with the healthcare team’. n.a. not determined (patients did not answer the question).

With regard to the patient-caregiver relationship, we wanted to know whether they feared a dehumanization of medicine, as there would be no tactile (or olfactory) contact, or at least a deterioration in the patient-caregiver relationship. We had 3 articles on this topic: Regarding the relationship with the healthcare team:

- Telemedicine dehumanizes medicine.
- Telemedicine weakens the relationship with carers.
- Monthly telemedicine makes it possible to maintain a strong link with the healthcare team. Patients’ responses were inconsistent and even contradictory, showing that they find it difficult to imagine the potential and procedures of teleconsultation before they have tried it. In fact, while 36% of respondents (24 respondents) said that they thought telemedicine dehumanized medicine (Fig 2D), 70% thought that it did not weaken relationships with healthcare professionals (Fig 2E) and even 93% of respondents (29 respondents) thought that telemedicine enabled them to maintain a strong link with the healthcare team (Fig 2F).

We asked patients if they thought that teleconsultation would allow them to avoid contamination with pathogens (such as the Covid19 virus) when they came for a consultation. The vast majority of patients did not answer this question. This highlights the lack of interest in this question and therefore possibly the low level of patient concern about this issue.

Finally, we looked at the practical aspects of teleconsultation. We started by assessing the fears that patients might have about the way teleconsultations are conducted. For example, we asked if they had any concerns about their ability to connect (digital literacy) and if they were worried about not being able to hear what the doctors were saying properly. 93% of patients had no concerns about their ability to connect, although only 70% said they did not need help with digital tools (Figs 3A and 3B). 100% of patients who answered this question said they were not worried about not hearing or understanding what the doctor was saying (Fig 3C).

**Figure 3:**
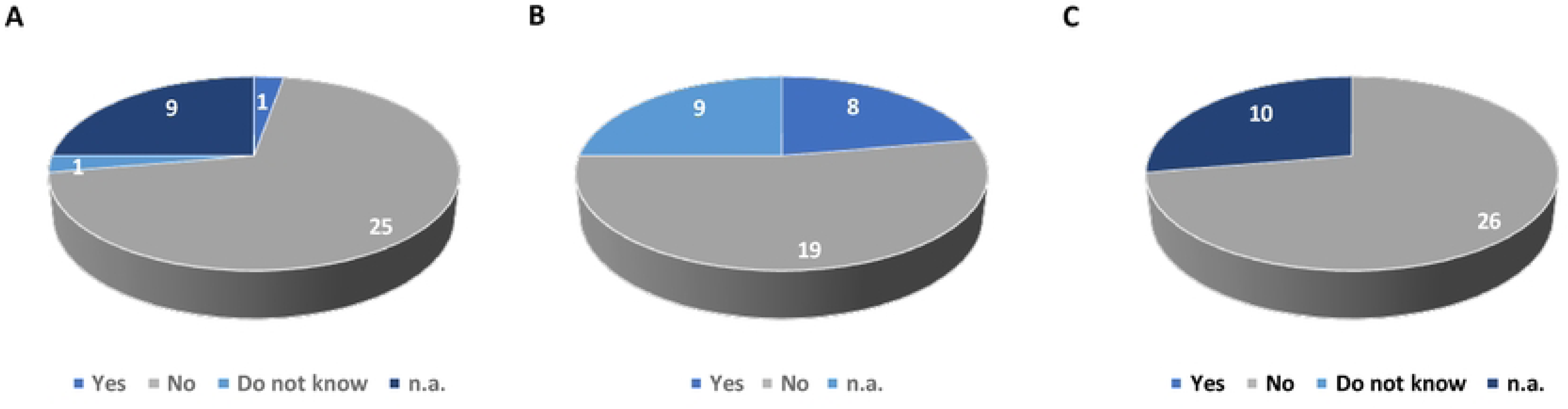
Patients’ potential fears about their ability to use digital tools A: Patients’ responses to the item ’I’m afraid I don’t understand how to get online’. B: Patients’ responses to the item: “I would prefer to have someone to help me to log in”. C: Responses to the question ’I fear I won’t be able to hear (comprehend) what the doctor will tell me. n.a. not determined (patients did not answer the question)

The practical aspects of teleconsultation were then considered. We started by assessing the fears that patients might have about the teleconsultation process. For example, we asked if they had any concerns about their ability to connect (digital literacy) and if they were worried about not being able to hear what the doctors were saying properly. 93% of patients had no fears about their ability to connect, although only 70% said they did not need help with digital tools (Figs 3A and 3B). 100% of patients who answered this question said they were not worried about not hearing or understanding what the doctor was saying (Fig 3C).

Finally, we asked patients if they were afraid that consultations would be recorded without their knowledge and for their personal use. None of the patients told us that they were worried about privacy, and none of them thought that their consultations could be recorded or listened to by anyone other than their carers. There is a high level of trust in the medical profession.

We found no correlation between gender, place of residence, level of education, occupation and opinion (positive or negative) of teleconsultation.

There was a very slight correlation between the age of the participants and their apprehension about consulting a doctor via a computer screen. It is the oldest patients who express some concern.

In conclusion, it is evident that the proposition of monthly telemedicine consultations has been met with a favourable response. There was a notable absence of any significant opposition, and patients have expressed a generally positive sentiment towards this novel approach to medical consultations, despite the recognition that it may not address all their medical concerns. It is important to note that during the enrolment process, participants were informed that their involvement in the study did not preclude the option of contacting the care team at any time or visiting the center if their general condition deteriorated. Telephone consultations were presented as an additional benefit in the follow-up.

### The consultation process

43 teleconsultations were conducted during the project. Overall, patients had no problems connecting. Most did so using a computer. There were no delays in connection.

The duration of teleconsultations ranged from 20 to 35 minutes.

Overall, the teleconsultations resulted in the prescription of medication. The prescriptions were e-mailed either to the patients’ homes or to their pharmacies.

These treatments may have prevented patients from needing hospitalization for their symptoms or made it harder for them to stay at home, as well as improving their symptoms and quality of life.

In addition, these systematic teleconsultations have made it possible to anticipate the implementation of home care coordination, such as HAH, to enable patients to remain at home and avoid unwanted hospital admissions. When certain symptoms (pain, vomiting, etc.) or clinical features (tumor wounds, general condition, etc.) deteriorate, more extensive medical care is required. In other cases, they have made it possible for the patient to be admitted directly to hospital (bypassing the emergency department) when it is not possible to provide care at home.

### Quantitative teleconsultation assessment

We then met the patients during their day hospital stay 3 months after their inclusion in the study. As mentioned previously [28], although one of the inclusion criteria was life expectancy > or equal to 3 months, only 13 of the 36 patients included in the study returned to the day hospital. The other patients died either before the first teleconsultation (11), between the first and second teleconsultation (7) or before the HDJ (Fig 4A). This highlights the fact that patients are referred to palliative care teams much too late in their care pathway. 44% had participated in at least one clinical trial during their illness. We found no association between clinical trial participation and 3-month survival.

**Figure 4:**
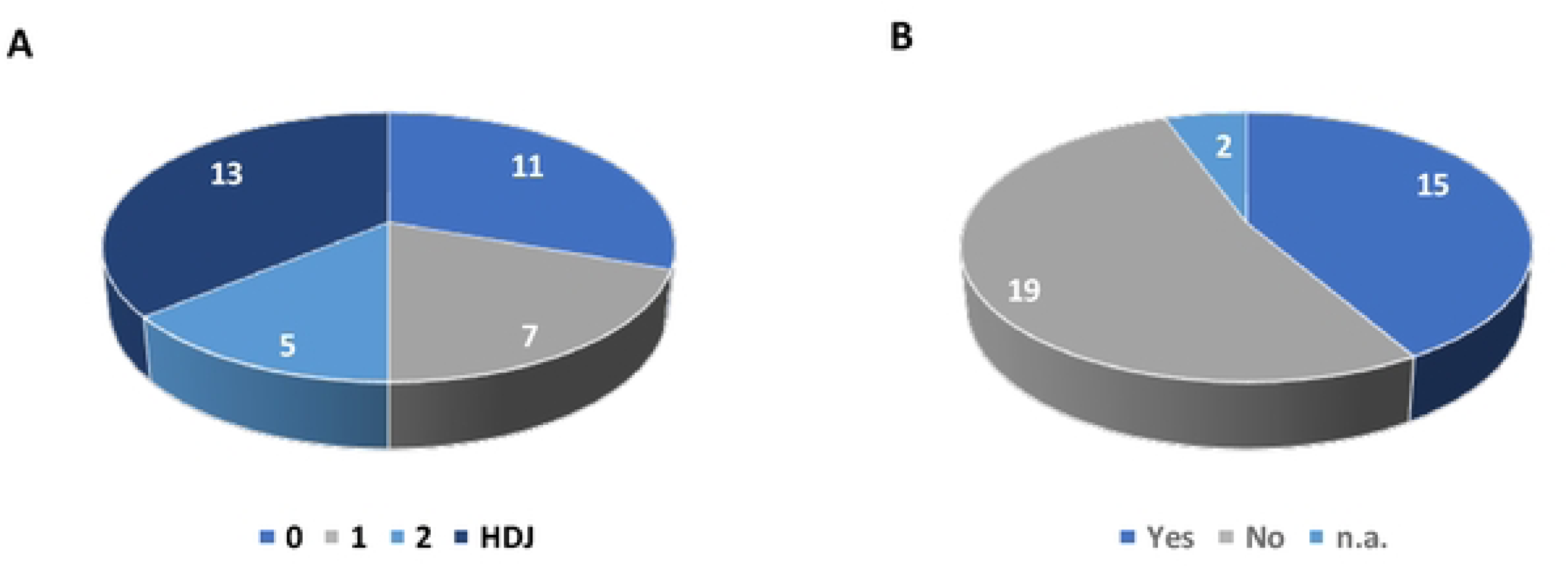
A: Distribution of patients according to whether they had 0, 1, 2 or 3 consultations. B: Number of patients who participated in a clinical trial before being referred to the to the palliative care department.

11 patients agreed to be interviewed about their experience of palliative care teleconsultation. Quantitative analysis of the interviews showed that 100% of respondents were satisfied with the teleconsultations. They mentioned the reassuring aspect of being able to see the EMSP doctor every month. 3 of them reiterated their reservations about the possible dehumanization of medicine. All of them said that they thought the teleconsultations were a plus for their follow-up. We asked them about the frequency of teleconsultations compared to face-to-face consultations. 64% thought that the frequency (2 teleconsultations for every face-to-face consultation) was optimal, while 36% thought that it would be better to alternate between teleconsultations and face-to-face consultations. No one mentioned the possibility of more teleconsultations.

### Qualitative analysis of patient’s freedom of speech in teleconsultations

#### 1/ Connectivity and mobility: a question of material and relational resources

We were interested both in understanding how teleconsultations can facilitate access to care in terms of geography, and in the material and relational dimensions that can act as barriers and/or levers. The patients interviewed highlighted the benefits of teleconsultation in terms of time savings and, above all, in terms of quality of life (less fatigue). Regarding time savings, we can quote the following sentences: “Yes, I’ve talked to my GP about it, for example, and I said it’s a pity he doesn’t do it either, because there are situations where you have to go to him, and there are other situations where it’s just small requests, small requests for letters or things like that, where it saves time both for them, for the professionals, and for us”.

Regarding the physical consequences of moving, which can lead to an increase in pain, we confirm that patients (or their carers) say that access to palliative care can be complicated by the physical wear and tear caused by the illness. I’m aware that sometimes you have to go to the doctor, but I find that it saves people like my mother, who is very tired, having to travel long distances, for example, to have a consultation like this to find out if everything is going well, it saves having to travel all the way from Toulouse again and tiring the person, teleconsultations like this are great, I think’.

So it’s true that not having to travel is important at the moment. Because I still have a lot of pain in my leg… well, my legs hurt. It’s true that I appreciate having less transport. But after that, coming in once in a while, as the protocol says… it’s also good to keep that visual, direct contact. We also have it in teleconsultation, but whatever.

Overall, patients were satisfied that the monthly teleconsultations enabled them to maintain medical follow-up without the fatigue or pain associated with travelling. We found no correlation between satisfaction with not having to travel and the distance from home to the care structure.

One of the aims of the project was to determine whether there was a specific profile of patients who could benefit from teleconsultation, with questions about age, social status and ease of use of digital tools.

As teleconsultations require appropriate computer equipment, they contribute to inequalities in access to healthcare by reproducing digital inequalities. Indeed, 2 patients could not be included in the protocol because they did not have access to digital tools. Also, although in the interviews all patients reported having a computer or telephone and a stable internet connection, access to technology is not determined by this factor alone. In other words, there may still be a difference between having the necessary computer equipment and practical knowledge of how to use it [29] ’The last time, what I didn’t understand was that I did it with the iPhone and I did the test, you know how to do it, I couldn’t see it or I could see it but I couldn’t hear it, there was something wrong, I didn’t understand why, especially as it was working, so my first consultation was by telephone. Then I took the tablet and managed to download Google, I don’t know what, and then with the tablet we managed to connect, but then the sound …, I had problems, I turned it up to the maximum, I had problems hearing […] lesson, maybe with a speaker, maybe more, should buy a Bluetooth speaker’.

While most people say that they can easily connect themselves (’No, because it’s really very easy, I think I also got the text message on my phone and the email to my email address, so all I had to do was click on the link and it would take us straight to the page’), the qualitative analyses show that mediation by a third party is often involved. Indeed, for patients with low computer skills, having someone close to them to facilitate the connection can be central to the teleconsultation process. As this interview extract shows, access to tele-consultation needs to be understood in relational terms (the patient’s relational resources) and in temporal terms (the moment of connection). It’s going well. You need someone who knows how to use the computer.

Researcher: But there’s Mister. And to be equipped. Patient : I couldn’t have done it alone.

Researcher : Maybe the nurse would have helped you.

Patient : Maybe. But given the time of the appointment, the nurse has to be there.

For this patient, her partner is an important resource, enabling her to carry out teleconsultations. Another uses her daughter to connect to the platform. When asked if it was easy for her to connect, she replied, “Well, you’d have to ask my daughter.

Thus, in addition to the material and practical conditions associated with digital technology, the social or relational capital of patients’ needs to be taken into account when setting up teleconsultations. However, the relational dimension, which can be mobilized as a resource through the use of a mediator with the necessary technological skills, can also be a constraint. In this case, access to the consultation depends on the availability of these third parties. The role of carers is therefore central to access to care and the evaluation of telehealth applications. Yes, it’s planned, that’s what’s planned. After that it’s the timetable, because my job is undergoing a lot of changes because my company is being sold and I don’t know who’s going to stay on, and since we’re in coaches, it’s special, (…) the employer who takes us over, we don’t know the timetable, what to do, it’s quite complicated. So what I said to her and explained to her, I said I don’t know if I will be available or not, because I need to be with her to connect and all that, so that she is not alone”.

Finally, the need for a third party to be present raises the question of the privacy of the exchange between doctor and patient. Firstly, the presence of carers for the teleconsultation raises the question of whether they remain in the same room to support the patient. Secondly, while most of the patients interviewed were able to isolate themselves at home for the teleconsultation (’my husband was there, but I was in my office’), we can assume that this is not necessarily the case for patients in certain family and social configurations (dense households, single-parent families, young children, etc.). These conditions may affect the transmission of certain important information for supportive care follow-up by patients, firstly because of the distraction that may be caused by third parties, and secondly because of the restricted freedom of speech.

#### 2/ Teleconsultation. A maximized route for “sentinel” patients

The Telemsos project was initially designed to include an initial face-to-face meeting, not only for purely medical reasons relating to the clinical examination and the various diagnoses, but also to facilitate the ’relational’ transition to teleconsultation.

In this way, patients meet the palliative care doctor only once at the HDJ before the teleconsultation is proposed. In this configuration, the phase of getting to know each other and its conditions are reduced in terms of the establishment of a relationship of trust. However, the majority of patients agreed to subsequent consultations by videoconference. According to the interviewees, these first face-to-face meetings, however limited, made it easier to move on to teleconsultation: ’The fact that I had met her once meant that I had a confident face (…) I felt I could trust her straight away’.

Because it’s a question of trust and then the fact is that if, the fact of having met Dr X before, of having spoken to her before, I think we’ve seen each other before’.

This adaptation of patients to teleconsultation, despite having less time to get to know the nurse, can be explained by the point at which they become part of the care pathway. The fact is that a transition to palliative care requires prior experience of the disease and its management. In the interviews, teleconsultation takes place at a time when a network of relationships with medical staff has already been established. In fact, palliative care concerns people who have relapsed and/or are considered to be in a chronic oncological situation. In general, these are people who have acquired experience of their disease and the technical and professional environment in which they are cared for (expert patients). In addition, some of them have already participated in other clinical trials (Fig 4B). These various factors make it possible to situate them in the context of what Anselm Strauss (1992) [30] calls an ’illness trajectory’, which takes on a professional dimension (’patient’s career’) corresponding both to ’the physiological evolution of the illness and to the whole organisation of the work deployed to follow this trajectory by all those involved in care, including the patient himself’. For patients and their relatives, it is a social experience of the physical manifestations of the illness (discomfort, fatigue, pain, etc.), as well as of the interactions with health professionals and the organization and coordination of the various actors involved. Teleconsultations thus appear to be an additional resource in their trajectory and in their own “work” as patients. Some patients even suggest the possibility of using teleconsultations not specifically for therapeutic purposes, but rather for ’care coordination’: Ah yes, yes, it’s in the same process, perhaps in a slightly more collegial way, we could see several people together, well, maybe it could be interesting to meet, for example, from the moment we have, how shall I say, if we can meet, as I already knew Doctor X, as I knew Doctor Y, (…)… why not continue with teleconsultations? I think it seems to me that we’re in a very sensitive area, of trust, and I think it could even be a slightly collegial meeting, but without being too intrusive’. In addition, some discussions took place in the presence of a professional, for example with the private nurse during a dressing change, which allowed the dressing protocol to be changed.

What’s more, for some patients who have a good command of the tools of telecommunications, these exchanges can make their medical journey a smoother one. This is particularly true when it comes to prescriptions: ’Yes, she also sent them directly to the pharmacy, there was a need for certain prescriptions that were encrypted, that’s all’.

For some people, these experiences of telemedicine in palliative care also form part of what might be called a ’patient’s telemedicine career’, which is built up alongside face-to-face consultations, but also forms part of a patient’s career. Most of the patients interviewed were having, or had already had, other teleconsultations. In this way, the use of digital tools in healthcare creates a new form of homecare for patients[31].

I like it because it’s difficult for me to move around now with my illness, so it allows me to overlap, one out of two consultations I do by teleconsultation with my GP”. I already use it for my husband. Because my mutual insurance company offers it at weekends and sometimes I need it for my husband at weekends. And I’ve used teleconsultation two or three times now. And every time it’s good’. In other words, the social characteristics of the patients interviewed make it possible to include them in the model of the “sentinel patient”, i.e. a patient capable of organising his or her own care and developing self-monitoring. This term was proposed by Patrice Pinell (1992)[32] in the context of cancerology to describe a person who is capable of taking a clinical look at his or her own body and of informing the doctor as soon as a symptom heralding the onset or recurrence of a disease is detected. Telemedicine therefore presupposes that patients have the ability to provide ’information’ to the doctor, to describe symptoms and physiological states, an ability that is socially situated (in particular according to gender and social class).

Ah, well, when I described my pain and everything to her… well, because I had a broken bone, a broken femur, she (the doctor) said to me, ’if you’re suffering like that, it’s not normal pain and I think there’s still something in the bone that’s causing you to suffer’. And it’s true that the bone was broken again. Because I’d already had a first operation and I’d had a gamma pin and the gamma pin was broken… because I also had metastases (…) She said to me during the consultation ’at the moment, are you in pain? So I stood up, we stood up together and I said ’there it is’. And that’s what it was. I owe her a debt of gratitude.

Another interesting point that emerges from these extracts is the fact that teleconsultation is very often presented less as a ’real consultation’ than as an additional channel used to maintain the relationship with the doctor: ’I like it because it’s difficult for me to travel now with my illness, so it allows me to overlap, one out of two consultations I do with my GP via teleconsultation’.

Teleconsultations are thus seen by patients as a resource for building relationships with professionals. The fluidity of telemedicine exchanges should therefore be seen as part of the continuity of therapeutic relationships that are already fluid in face-to-face settings. Above all, however, it must be socially situated. In other words, this ’one-to-one’ discussion between patient and doctor, which involves ’the spoken word’ (Memmi, 2003), is a mode of intervention that favours those with medical cultural capital - in other words, those who are deemed able to express themselves verbally without challenging medical authority. Ultimately, teleconsultation reflects the same sociological issues as face-to-face consultations. As social science research shows, care is often effective with patients who follow medical norms, who like to be understood, who ask enough questions to keep track of their treatment, but who do not challenge medical authority. It is these patients who seem to be the focus of Telemsos’ research. For them, these teleconsultations represent an opportunity in the treatment process. In fact, they are part of a trajectory rooted in the ’careers of the sick’. They therefore have a fairly high level of cultural and medical capital, in other words a high level of health literacy, and actively position themselves as ’sentinels’ for their own health.

#### 3/ Giving substance to images and words

We also wondered how easy it would be for patients to talk to their doctor during a teleconsultation, in line with our concern to determine whether there was a patient profile suitable for teleconsultation. The issues of doctor-patient interaction need to be considered in the light of the patient’s social background, as the ability to verbalize one’s own body and bodily sensations is socially constructed and situated, and varies with levels of health literacy. People with ’low’ literacy have more difficulty describing their symptoms, expressing their concerns and managing their illness [33][34].

The people we interviewed had educational and cultural capital, which is strongly linked to health literacy, built up over long careers as patients. We can therefore observe a certain fluidity in the exchanges during teleconsultations:

’Researcher: Were you embarrassed to ask questions? No. I’m telling you, the relationship has grown stronger!

Researcher: Is that because of the personality of the (doctor) or because of the screen?

A bit of both, I think. I’d say both, because I’m easy to talk to and so is Madame X and, well, she hears everything we say. To get back to what she felt.

While some physical signs are easier to demonstrate or show (’Yes, I was able to show her too, my tumour wound for example, there you go’), other signs are more complex to show or imitate and the exercise requires patients to be particularly performative. Expressing symptoms or questions requires the ability to put the body and its sensations into words. One woman commented on the differences between face-to-face and video consultations, in particular the difficulty of expressing bodily signs verbally as they are invisible to the camera: It wouldn’t have been easy. Because it’s not very big and it’s skin-coloured. They’re little circles. But you can take a blood sample beforehand… but measuring your heart rate, for example… So there are differences. I would say that teleconsultation is a good way of compensating for that, but it’s not as good as a real consultation.

Some of the patients interviewed also expressed the need for the teleconsultation to take a long time to obtain information and/or to verbalize the different physical sensations they experienced.

I found that almost 20 minutes was too short […] I think it was both of us, at the end of 20 minutes, it seems to me that we said 20 minutes and so we stopped at 20 minutes […] maybe even longer, in the sense that I thought maybe a little bit longer’.

Although patients say that it is possible to demonstrate (’part of my body (that) needed to be seen!’), visualizing the body requires them to be performative and to adopt an attitude that needs to be even more active in teleconsultation: ’So I got up, we got up together and I said “there it is (the pain)”’. Once again, it is important to situate these patients socially, as they have educational and medical capital and say they feel comfortable explaining their feelings to doctors via a screen. When asked if they found it difficult to explain their health concerns, the majority said no, as this patient put it: ’No, not at all, because we have a face and we can express what we want’.

Again, we need to return to gender differences in therapeutic relationships, whether face-to-face or remote, and how they relate to patients’ social class. Relationships with the body and with pain differ by gender [35,36], as do the relationships that men and women have with the medical world (professionals, medical products, etc.) beyond their social class, particularly in the case of cancer [37]. Gender differences also emerge in the way people participate in supportive care, particularly in telemedicine consultations, where the issues of ’being silent’, ’saying’, ’showing’ and ’doing’ are not combined in the same way. The research conducted by Dudoit et al (2007) [38] with patients in oncology support care shows that women tend to ’put themselves and their bodies to work’, which allows and facilitates touching and talking about their own feelings. For men, closer to the idea of a body as an object rather than a body as a lived experience, the dominant model is to ’make their bodies available’. In line with the norms of masculinity, they play little role in mediating between the body and the ’discourse’ about the body. Since these forms of mediation are all the more necessary as the consultation takes place through the interface of a screen, the results of the analysis must be considered in relation to the profiles of the interviewees, all of whom are women and therefore fall within the first model of participation in care.

## DISCUSSION

In the context of the present high demand for palliative care and the consequent shortage of available beds in care units, teleconsultations appear to be a potential aid in the monitoring of cancer patients. It is important to note that their intention is not to replace face-to-face consultations, but rather to provide additional support and to maintain a strong link between the hospital care team and the patient.

The objective of the present study was to ascertain whether teleconsultation follow-up could be made available to all patients, irrespective of gender, age, digital literacy, social class, geographical location, and preconceptions about telemedicine. The participants in this study were attending the EMSP for the first time when teleconsultation was offered. The objective was to ascertain whether the requisite trust for telemedicine follow-up could be established in a single face-to-face consultation. Patients newly referred to the EMSP are typically at an advanced stage in a curative project or clinical trial and are often anxious about the progression of their pathology. However, they are also likely to be well-informed about their care pathway, which has frequently been protracted.

The majority (80%) of patients recruited to the trial were women, yet we are unable to attribute this discrepancy in participation between the sexes to life expectancy in France or to recruitment bias, as all patients approached consented to participate. It is noteworthy that the institute where the study was conducted has a high rate of individuals diagnosed with breast cancer, and that the population under study is predominantly from affluent social backgrounds. The cost of care, including palliative care, is covered by the state for all citizens in France, and thus we did not consider income to be a factor in the recruitment bias. However, we can hypothesize that gender or social level may influence the longevity of patients in oncological care, with women from more comfortable social backgrounds potentially better able to cope with the constraints of an oncology treatment plan than their male counterparts. It has also been reported that people with very low incomes have a lower life expectancy than others, which may explain their limited access to the palliative care team. The introduction of teleconsultation was intended to reduce inequalities in access to care; however, two out of 38 patients did not have access to a connection and could not be recruited, indicating that approximately 5% of patients could not benefit from teleconsultations in addition to HDJ palliative care.

In terms of preconceived notions regarding telehealth, it was observed that patients expressed a favorable opinion of this type of remote monitoring, despite a lack of prior experience. Indeed, 100% of patients who were offered monthly teleconsultations and had a domestic connection accepted the offer. Concerning preconceived notions at the time of enrolment, patients reported minimal trepidation. They acknowledged the advantages of monthly monitoring and the convenience of avoiding travel. However, 60% of patients expressed the misconception that teleconsultation would be applicable only to specific medical procedures. Upon enrolment, no association was identified between gender, place of residence, level of education or occupation, and an a priori favorable opinion of teleconsultation. Older patients exhibited minimal reluctance, largely attributable to concerns regarding their capacity to utilize digital tools. No patient reported concerns regarding privacy.

The study set out to ascertain whether the uptake and feasibility of teleconsultation for palliative care patients could be influenced by age, gender, socioeconomic level or pathology. The findings revealed that these factors did not influence the success of teleconsultation. In fact, no major difficulties were encountered by patients in connecting, regardless of their age. Some were accompanied to the teleconsultation. The benefits of teleconsultation for the clinical and psychological management of patients were evaluated, and the value of monthly follow-ups was demonstrated. All patients reported benefit from the consultations, and a relationship of trust with their doctor was easily established, despite their lack of knowledge prior to the teleconsultation. The teleconsultations, like the face-to-face consultations, helped to highlight deteriorating medical situations or changes in quality of life. The doctor was able to make a diagnosis and propose a treatment plan (prescriptions sent by e-mail, liaison with the HAH).

The patients expressed a desire to continue with monthly teleconsultation follow-ups following their three-month participation in the study. In the absence of the possibility of monthly face-to-face consultations, the value of developing teleconsultations for patients undergoing palliative care in oncology is demonstrated. Telemedicine in palliative care facilitates the monitoring of a larger number of patients and improves equity of care.

However, it is also demonstrated that patients are referred to the mobile palliative care team too late. A significant proportion of the patients who were offered telemedicine follow-up (64%) passed away within three months of enrolment, indicating a delay in accessing palliative care. This underscores the necessity for palliative care to extend beyond the end-of-life stage and emphasizes the importance of promoting both telemedicine and early access to palliative care for patients.

The sociological analysis explores multiple dimensions; however, three aspects are pivotal to the demonstration:

- It is imperative to shift the prevailing definition of ’access to care’ from its current geographical and spatial context. A comprehensive analysis of interview data reveals that the concept of access, in its traditional sense as mobility, must be redefined to encompass a more comprehensive understanding of connectivity. This shift necessitates a re-evaluation of both geographical and ’remote’ access, emphasizing the role of material, technical, physical, temporal, and relational factors.
- It is imperative to understand teleconsultation not as an isolated practice, but as a resource that contributes to the array of tools professionals and patients utilize to navigate the journey through illness.
- The integration of this resource into a patient’s pathway is socially situated and contingent on specific social profiles with high levels of educational, cultural and medical capital.
- Finally, an in-depth analysis of the content of teleconsultations and the doctor-patient relationship, as well as the ability of the patients interviewed to perform their bodies and their discourse, once again demonstrates the importance of educational, cultural and medical capital for the success of teleconsultations. It should be noted that gendered socialization also plays a role in the construction of these differentiated patient models.

Consequently, the present project has facilitated patient access to monthly teleconsultations for follow-up with the palliative care department.

All authors have read the manuscript and declare that they have no conflicts of interest.

We thank the Association pour la Recherche contre le Cancer (ARC) for funding this study.

## Data Availability

All relevant data are within the manuscript and its Supporting Information files.

## Acknowledgments

We thank the Association pour la Recherche contre le Cancer (ARC) for funding this study.

